# Covid-19 and Inequity: A comparative spatial analysis of New York City and Chicago hot spots

**DOI:** 10.1101/2020.04.21.20074468

**Authors:** Andrew Maroko, Denis Nash, Brian Pavilonis

## Abstract

There have been numerous reports that the impact of the ongoing COVID-19 epidemic has disproportionately impacted traditionally vulnerable communities, including well-researched social determinants of health, such as racial and ethnic minorities, migrants, and the economically challenged. The goal of this ecological cross-sectional study is to examine the demographic and economic nature of spatial hot and cold spots of SARS-CoV-2 rates in New York City and Chicago as of April 13, 2020.

In both cities, cold spots (clusters of low SARS-CoV-2 rate ZIP code tabulation areas) demonstrated typical protective factors associated with the social determinants of health and the ability to social distance. These neighborhoods tended to be wealthier, have higher educational attainment, higher proportions of non-Hispanic white residents, and more workers in managerial occupations. Hot spots (clusters of high SARS-CoV-2 rate ZIP code tabulation areas) also had similarities, such as lower rates of college graduates and higher proportions of people of color. It also appears to be larger households (more people per household), rather than overall population density, that may to be a more strongly associated with hot spots.

Findings suggest important differences between the cities’ hot spots as well. They can be generalized by describing the NYC hot spots as working-class and middle-income communities, perhaps indicative of service workers and other occupations (including those classified as “essential services” during the pandemic) that may not require a college degree but pay wages above poverty levels. Chicago’s hot spot neighborhoods, on the other hand, are among the city’s most vulnerable, low-income neighborhoods with extremely high rates of poverty, unemployment, and non-Hispanic Black residents.

## Introduction

There have been numerous reports that the impact of the ongoing COVID-19 epidemic has disproportionately impacted traditionally vulnerable communities, including well-researched social determinants of health(1), such as racial and ethnic minorities, migrants, and the economically challenged both globally (2) and domestically (3). For instance, according to an April 8^th^ Centers for Disease Control and Prevention’s Morbidity and Mortality Weekly Report, of the 580 hospitalized patients in 99 counties in 14 states found that non-Hispanic (NH) black were disproportionately affected (4). Poverty and low income have also been identified as important issues not just as a function of potentially classist policy, but also due to the increased difficulty for those in more tenuous financial situations and occupations to implement physical distancing and isolation. Individuals at or below the poverty line are affected by residential overcrowding, increased smoking rates (5), exposures to environmental pollutants (6), and lack of access to healthcare according to a UN report - all of which can increase the spread of the virus or cause adverse outcomes (7). However, it is important to note that at this point there is very little data at smaller geographies (e.g., counties or sub-county units) that report on demographic or economic characteristics of those with the virus, hospitalizations, or deaths (8).

These types of health disparities, which are not new to this pandemic, may be a function of elements of the social and physical environments, as well as factors associated with systemic and institutionalized racism and classism resulting in suboptimal access to resources, including health care and support services (9, 10). Additionally, these vulnerable communities’ risk of having worse outcomes from COVID-19 infection may be increased due to a higher prevalence of comorbidities or underlying conditions such as asthma, diabetes, and other conditions (11-14). This suggests that existing health disparities are likely to be magnified in the context of COVID-19, and potentially extend well beyond the lifespan of the epidemic due to sociodemographic inequity, and economic hardships at the global, national, and local levels which themselves have a differential impact on individuals and populations both directly and indirectly (15-18).

Scale is an important element when examining any phenomenon that has a locational component, and the choice of both unit of analysis and study area can have large effects on the outcomes of the analyses. For instance, when quantifying the association between a health outcome and population density, using a county-scale could obscure the heterogeneity of both the outcome of interest (e.g., there may be very high rates in one part of the county, and low rates everywhere else) and the associated variable (e.g., one neighborhood in the county could house a large number of resident in high-rise buildings and the remainder could be low intensity single family homes and park land). This issue, often called the modifiable area unit problem (MAUP), has been documented in multiple research domains, including health (19, 20), exposure estimation (21), measures of access (22), and environmental justice (23, 24). A related phenomenon is the choice of the overall study area. For instance, based on the American Community Survey 2018 5-year estimates (25), Staten Island, one of the boroughs of New York City, may seem to have very high population density (over 8000 people per square mile) when compared other counties in the entire U.S., some of which have values far below one person per square mile (e.g., Denali, Alaska and Esmeralda, Texas). However, Staten Island is the least densely population borough of New York City, with Manhattan having more than 70,000 residents per square mile. As such, it would appear comparatively dense at a national scale, but relatively less-dense at a city scale. Since social interaction is the basis of community transmission of the virus, it stands to reason that denser areas would be more rapidly and severely impacted than low-population areas.

Although this may be true at a national or even global level at this stage of the epidemic (26), where transmission may be easier in urban regions versus rural areas, it is unclear if this is the case at an intra-urban, sub-city, scale.

The understanding of the disease characteristics at national and global scales is unquestionably important, however it is also necessary to appreciate its dynamics at a more granular, neighborhood-level. According to data collected by the New York Times aggregated by county, as of April 13, NYC has the most reported cumulative cases (106,764) and deaths (7,154) in the United States. Cook County, Illinois, which contains Chicago, is fifth on the list for that day, with 15,474 cases and 543 deaths (27). However, the spatial distribution of reported positive cases are not homogeneously distributed across either city.

The goal of this ecological cross-sectional study is to examine the spatial and demographic nature of reported SARS-CoV-2 diagnoses in New York City (NYC) and Chicago (CHI) as of April 13, 2020.

Specifically, we examine SARS-CoV-2 diagnosis rates per ZIP code tabulation area (ZCTA) and compare sociodemographic and economic characteristics between spatial hot spots and cold spots. The characteristics of the NYC and CHI hot / cold spots are then compared to reveal differences and similarities between the cities.

### Data

Cumulative counts of SARS-CoV-2 diagnoses (“cases”) for NYC are from New York City Department of Health and Mental Hygiene’s Incident Command System for COVID-19 Response (4/13/2020) (28). Of the 211 Zip Code tabulation areas in NYC, 34 (16.1%) had no data, likely due to low or no populations (e.g., airport, commercial areas, parks). Reported SARS-CoV-2 diagnoses (> 5 cases per ZIP code) are from the Illinois Department of Public Health (4/13/2020) (29). ZCTAs were included in the CHI analysis if their centroid was within the published city boundary or greater than 20% of the ZCTA area was within the city. Of the 60 ZCTAs, three (5.0%) were excluded due to missing data, likely due to low populations or reporting five or fewer cases. Demographic and economic data are from American Community Survey (ACS) 2018 5-year estimates via NHGIS.org (25).

It is vitally important to note that these data may not necessarily represent the true distribution of SARS-CoV-2 based on biases in testing and extremely limited or differential testing and/or access. Testing for SARS-CoV-2 also has resulted in high percentage of false negatives. This is due to two reasons, development of rapid testing during an epidemic and need the need to collect a sample from deep in the pharynx which can be uncomfortable for the patient and is time consuming for the health care worker. False negatives are especially problematic since the infected individual can continue to spread the virus without being aware. There have also been instances of false positives being reported in health care setting due to the high viral load present in the environment (30). These factors can decrease the reliability and accuracy of the data and make comparisons within and across geographical regions difficult.

## Methods

Raw rates of reported SARS-CoV-2 cases were calculated by normalizing the number of diagnoses by the ZCTA population (cases per 1,000 residents) and spatialized and analyzed using ArcGIS 10.7 (ESRI, Redlands California). The Global Moran’s I based on ZCTA contiguity revealed clustering (z-scores = 5.14 and 13.2 for CHI and NYC, respectively. Both p-values < 0.001), suggesting that it is highly unlikely for the clustered pattern to be a result of random chance. Hot spots of rates for each city were then calculated using the Getis-Ord (GI*) statistic parameterized using contiguity (i.e., ZCTAs sharing a boundary or corner). Resulting hot spots represent clusters of contiguous ZCTAs with higher values within the city (GI* was calculated once for NYC and once for CHI), whereas cold spots represent clusters of ZCTAs with low values. Clusters with >= 95% confidence were included in the analyses (*figure 1)*. It is important to note that identifying statistical hot and cold spots is not the same as simply selecting the ZCTAs with the highest rates (e.g., top quartile). Unlike using quantiles or some other classification technique, hot (or cold) spots require clusters of ZCTAs to have high (or low) values relative to the study area which are accepted or rejected based on a significance value.

**Figure 1:**
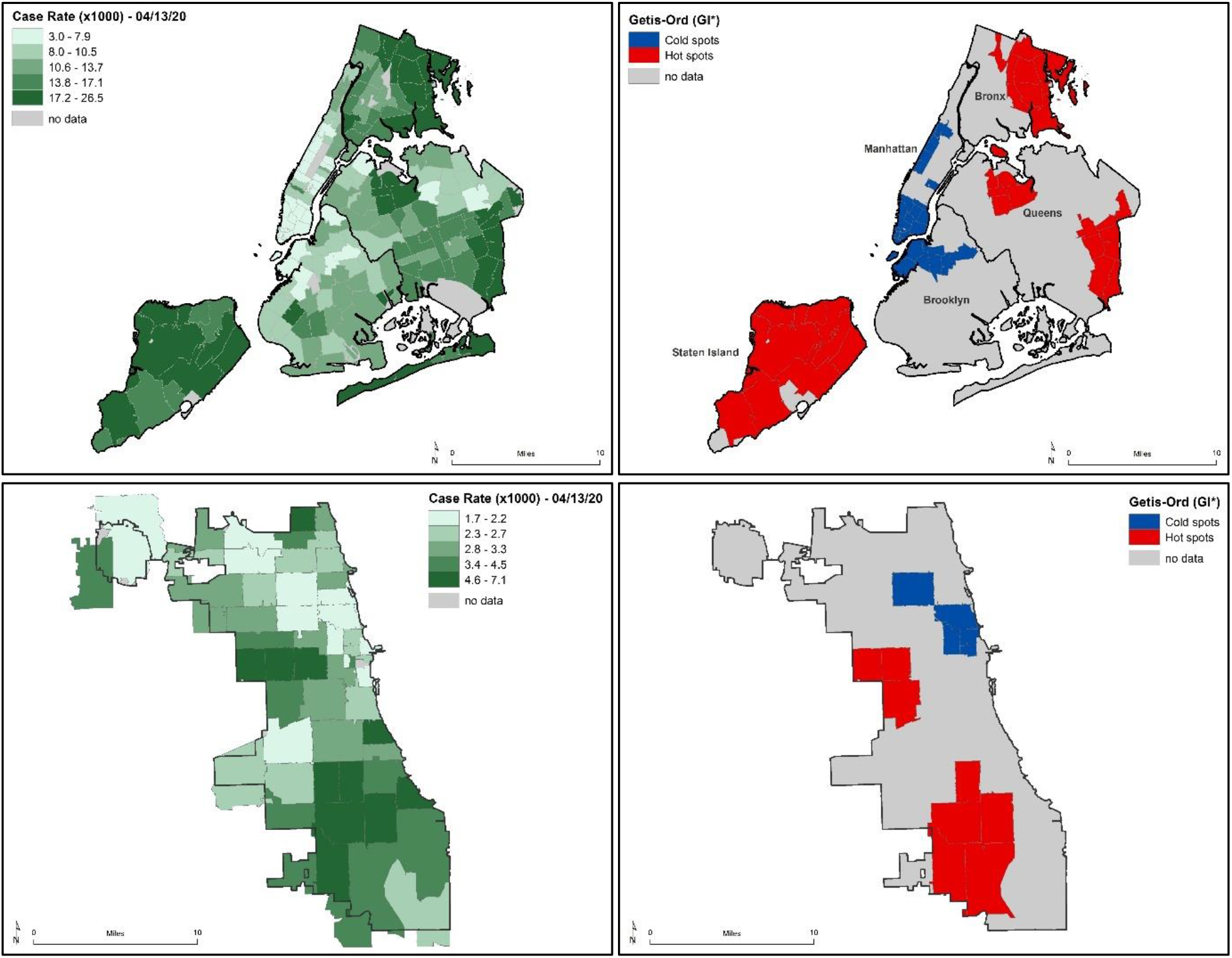
Reported SARS-COV-2 cases per 1,000 ZCTA residents. Note: cities are shown at different scales. TOP LEFT: NYC raw rates in quintiles. TOP RIGHT: NYC hot spots and cold spots (GI*) at >= 95%, confidence. BOTTOM LEFT: CHI raw rates in quintiles. BOTTOM RIGHT: CHI hot spots and cold spots (GI*) at >= 95%, confidence.

American Community Survey (ACS) data were mapped by ZCTA and linked with SARS-CoV-2 case rates and hot/cold spots for exploration and analysis (Figure 2). ZCTA averages, stratified by hot/cold spot status, were calculated for variables of interest which include: (1) SARS-CoV-2 cases per 1000 residents, (2) total population, (3) population density, (4) average household size, (5) % of housing units with > 1 occupant per room, (6) % NH White, (7) % NH Black, (8) % Latinx / Hispanic, (9) % foreign-born, (10) % 65 years or older (11) % of workers who commute using public transportation, (12) % of adults without a high school degree, (13) % of adults with a bachelor’s degree or higher, (14) % of residents earning under the federal poverty threshold, (15) median household income, (16) % of the civilian workforce who is unemployed, and (17) % in of workers in management, business, science, and arts occupations.

**Figure 2:**
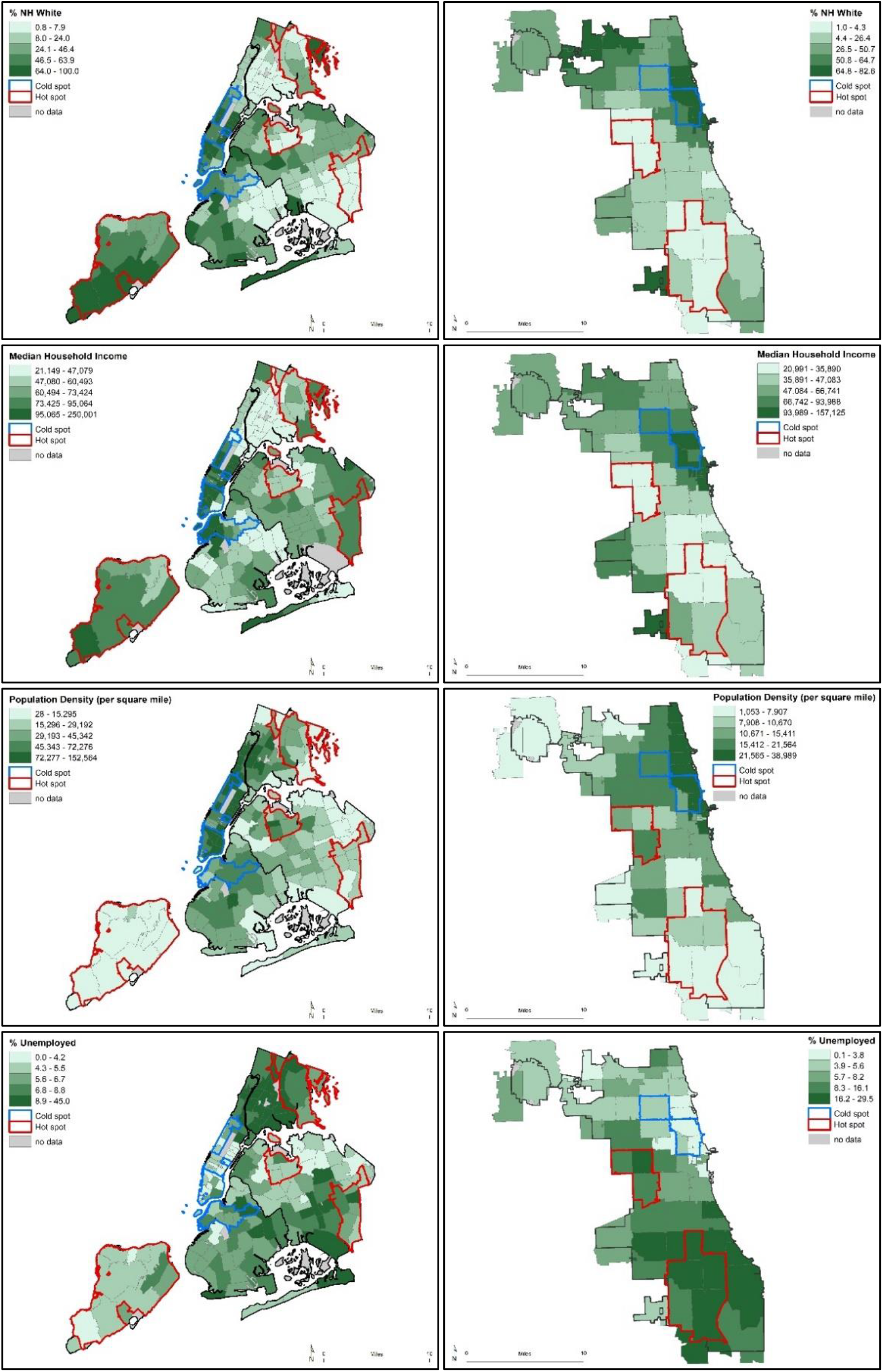
Selected ZCTA-level demographics for NYC and CHI in quintiles. Note: cities are shown at different scales. TOP ROW: % NH white. SECOND ROW: median household income (in 2018-adjusted dollars). THIRD ROW: population density (people per square mile). FOURTH ROW: % unemployed.

SAS Statistical software (version 9.4, SAS Institute, NC) was used to conduct all statistical analyses. Means were calculated across ZCTA for both cities. A Wilcoxon Two-Sample Test was used to calculate differences in demographic variables at the ZCTA level between the two cities. Due to the small sample size the t-approximation was used to calculate p-values.

## Results

Demographics in the NYC and CHI study areas are summarized and described using population-weighted averages (*table 1*). The overall positive test rate on April 13 for NYC was 12.5, which was approximately four times greater than CHI. New York City and CHI were comparable in terms of median household income, percent non-Hispanic White, percent with a bachelor’s degree, and percent below the poverty line. The main difference between the two cities was in terms of population density and percent foreign-born. Although NYC and CHI are roughly the same area (294 mi^2^ and 262 mi^2^, respectively) NYC has almost three times as many people per mi^2^. The density is also observed in households, with NYC having nearly three times the rate of having more than one person per room compared to CHI. The dissimilarities in density and urban landscape are also reflected in the percentage of individuals using public transportation, with 56.3% of NYC residents relying on public transportation and only 27.6% in CHI.

**Table 1:** NYC and Chicago city-wide demographics

| Variable | NYC* | CHI* |
| --- | --- | --- |
| # of ZCTAs | 211 | 60 |
| Case rate (per 1000) | 12.5 | 3.3 |
| Population (in millions) | 8.4 | 2.8 |
| Area (square miles) | 294 | 262 |
| Population density (in thousands per square mile) | 28.7 | 10.8 |
| Average household size | 2.7 | 2.6 |
| % >1 occupancy per room | 9.0 | 3.8 |
| % NH white | 32.1 | 33.1 |
| % NH Black | 22.0 | 29.4 |
| % Latinx/Hispanic | 29.1 | 29.1 |
| % foreign-born | 37.0 | 20.8 |
| % 65 year and older | 14.1 | 12.1 |
| % public transp. commuter | 56.3 | 27.6 |
| % without high school degree | 18.4 | 15.5 |
| % with bachelor's or higher | 37.4 | 37.8 |
| % in poverty | 18.9 | 19.3 |
| Median household income (in thousands of dollars) | 64.2 | 57.3 |
| % unemployed | 6.9 | 8.9 |
| % management occupation | 41.3 | 40.4 |

\* These data are based solely on ZCTAs used in the analysis and may differ from published demographic estimates as well as SARS-CoV-2 rates (as of 4/13/20).

Demographics in the NYC and CHI hotspots were summarized and described using ZCTA averages (*table 2*). There were striking differences between hot spots and cold spots in each city, as well as marked differences comparing across study areas. The NYC hot spots included 31 ZCTAs, representing nearly 1.5 million people (∼ 17.4% of the population in the NYC study area) and CHI hotspots consisted of 8 ZCTAs with 445,000 residents (∼15.8% of the CHI study area population). Hot spot neighborhoods in both cities tended to have lower proportions of non-Hispanic (NH) white residents, higher proportions of NH Black / African-American residents, a greater percentage of older residents, fewer college graduates, and lower proportions of workers in managerial occupations compared to cold spots or the “rest of city”.

**Table 2:** Hot / cold spot characteristics of NYC and Chicago (ZCTA-level averages)

| Variable | NYC |  |  |  | CHI |  |  |  |
| --- | --- | --- | --- | --- | --- | --- | --- | --- |
|  | cold spot | hot spot | rest of city* | hot vs. cold p-value | cold spot | hot spot | rest of city* | hot vs. cold p-value |
| # of ZCTAs | 28 | 31 | 118 | -- | 5 | 8 | 44 | -- |
| Case rate (per 1000) | 6.8 | 19.2 | 11.9 | -- | 2.2 | 4.9 | 3.1 | -- |
| Population (in millions) | 1.11 | 1.46 | 5.85 | -- | 0.24 | 0.45 | 2.13 | -- |
| Area (mi<sup>2</sup>) | 17 | 93 | 175 | -- | 12 | 48 | 202 | -- |
| Population density (in thousands per square mile) | 68.9 | 22.9 | 45.3 | <0.01 | 23.4 | 10.0 | 15.7 | 0.02 |
| Average household size | 2.1 | 3.0 | 2.7 | <0.01 | 2.0 | 2.8 | 2.5 | 0.03 |
| % >1 occupancy per room | 6.1 | 7.9 | 8.9 | 0.39 | 2.1 | 3.9 | 3.9 | 0.36 |
| % NH white | 57.0 | 27.1 | 33.6 | <0.01 | 66.1 | 4.3 | 40.1 | 0.01 |
| % NH Black | 11.4 | 29.5 | 19.1 | 0.01 | 7.2 | 82.9 | 23.0 | 0.01 |
| % Latinx/Hispanic | 12.9 | 28.9 | 28.4 | <0.01 | 15.7 | 11.4 | 25.8 | 0.12 |
| % foreign-born | 24.6 | 35.5 | 37.8 | <0.01 | 15.7 | 5.8 | 22.7 | 0.06 |
| % 65 year and older | 12.6 | 15.0 | 14.5 | 0.04 | 9.1 | 14.5 | 12.4 | 0.04 |
| % public transp. commuter | 58.4 | 42.1 | 55.1 | <0.01 | 32.5 | 30.1 | 23.9 | 0.62 |
| % without high school degree | 8.6 | 16.6 | 18.5 | <0.01 | 5.8 | 20.0 | 13.5 | 0.04 |
| % with bachelor's degree | 70.5 | 27.9 | 36.8 | <0.01 | 72.6 | 16.7 | 41.2 | 0.01 |
| % in poverty | 12.7 | 13.5 | 18.4 | 0.47 | 9.9 | 31.2 | 17.8 | 0.01 |
| Median household income (in thousands of dollars) | 117.3 | 70.0 | 64.3 | <0.01 | 96.5 | 34.6 | 63.1 | 0.01 |
| % unemployed | 4.8 | 6.5 | 7.1 | <0.01 | 3.7 | 17.7 | 8.6 | 0.01 |
| % management occupation | 67.1 | 33.6 | 39.7 | <0.01 | 62.7 | 23.1 | 41.2 | 0.01 |

\* "Rest of city" refers to ZCTAs that were not in hot or cold spots.

Spatial density is an important factor in the spread of communicable diseases. Hot spots in both cities had significantly larger household sizes compared to cold spots. However, hotspots were located in neighborhoods that were significantly less dense and the proportion of housing units with more than one occupant per room were not significantly different (0.39 and 0.36 in NYC and CHI, respectively) between hot and cold spots. Additionally, there were lower proportions of public transportation commuters in both cities’ hot spots than cold spots, the difference in NYC (p < .01) was more meaningful than that in CHI (p = 0.62). This is not reflective of public transportation use during the outbreak, but rather a measure of “connectedness” or “centrality” of a neighborhood.

There are some variables that suggest different patterns between NYC and CHI. Poverty rates, for instance, are lower for both hot and cold spots compared to the rest of the city in NYC; whereas in CHI the poverty rates are highest in the hot spots. Unemployment follows a similar trend, where the NYC rates are highest in the areas which are neither hot spots nor cold spots, but in CHI the rates are by far the highest in the hot spots. Although median household income is highest in cold spots for both cities, in NYC the income in hot spots is higher than the rest of the city, whereas in CHI hot spot incomes are much lower than the rest of the city. Finally, the proportions of both foreign-born (p < 0.01) and Latinx (p < 0.01) residents are higher in NYC hot spots than cold spots (but hot spot values are similar to the rest of the city), whereas the opposite is true for Chicago with foreign-born (p < 0 .06) and Latinx (p = 0.12) appearing protective.

## Conclusion and Discussion

In both Chicago and New York City, cold spots demonstrated typical protective factors associated with the social determinants of health and the ability to social distance. These neighborhoods tended to be wealthier, have higher educational attainment, higher proportions of non-Hispanic white residents, and more workers in managerial occupations. Hot spots between the cities also had some similarities, such as lower rates of college graduates and higher proportions of people of color. However, there are some other findings which must be highlighted. For instance, in both cities it is not the densest areas which appear to be most impacted by SARS-CoV-2, but rather it is the less-centralized, lower-density neighborhoods. In these two large U.S. cities it appears to be larger households (more people per household), rather than overcrowding or overall population density - which may be reflective of neighborhood socioeconomic status - that may be a more strongly associated with geographic hot spots.

Perhaps most striking are the differences in the economic and racial composition of the hot spots between NYC and CHI. At this point in the epidemic, NYC has a mix of racial/ethnic neighborhoods. For instance, the Staten Island hot spot in NYC is nearly 60% NH white, whereas the hot spot in Eastern Queens is less than 6% NH white (*figures 1 and 2*). In NYC overall, the ZCTA-level average shows approximately 27% of the population as NH white, 30% NH Black, and 29% Latinx/Hispanic. In Chicago, the hot spots ZCTAs are on average approximately only 4% NH white, 11% Latinx/Hispanic, and nearly 83% NH Black. Although in both cities NH white residents may be underrepresented in hot spots, Chicago shows the inequities much more clearly. Economic distinctions are even more stark. The population in NYC’s hot spots are, overall, middle income with ZCTA-level average median household incomes around $ 70,000, which, although lower than the cold spots ($ 117,000), are higher than the rest of the city ($ 64,000). Conversely, the average median household income in Chicago’s hot spots is only $ 35,000 with cold spots and the rest of the city being $ 97,000 and $ 63,000, respectively. Poverty rates in NYC hot and cold spots were both around 13%, whereas the rate in the rest of the city was over 18%. Chicago, on the other hand, had hot spots with poverty rates of over 30% which is higher than both cold spots (10%) and the rest of the city (18%). This is mirrored by unemployment rates, where the NYC hot spots had rates of under 7% compared to Chicago’s nearly 18%.

These characteristics can be generalized by describing the NYC hot spots as working-class and middle-income communities, perhaps indicative of service workers and other occupations (including those classified as “essential services” during the pandemic) that may not require a college degree but pay wages above poverty levels. Chicago’s hot spot neighborhoods are among the city’s most vulnerable, low-income neighborhoods with extremely high rates of poverty, unemployment, and NH Black residents.

It is important to note that this represents an ecological analysis and does not use individual level data. The results characterize the neighborhoods (clusters of ZCTAs) and not necessarily the individuals living in those neighborhoods. The goal of this project is not to suggest causation, but rather to demonstrate the nature of SARS-CoV-2 hot and cold spots in two large U.S. cities. The information about the demographic and economic characteristics of hardest hit areas may help direct resources to mitigate the impact of COVID-19 properly and preemptively. However, the differences found, although striking, may be at least partially a function of a number of factors, including potential bias and extremely limited testing/reporting and possible false positives/negatives. For instance, it is possible that the Staten Island cluster is a result of more aggressive testing practices in those neighborhoods compared to other areas with less social or political capital. It is also important to note that this analysis is based on testing results, and do not examine COVID-19-related hospitalizations or deaths. Additionally, NYC and CHI are not only different in urban morphology and demographics, but also may be in different stages of the epidemic. The cumulative SARS-CoV-2 rates, particularly when comparatively low as is the case in some Chicago ZCTAs, can change rapidly due to the dynamic nature of infectious disease spread as well as a hopeful increase in thorough testing. However, it is clear that as of April 13, 2020, Chicago and New York City have some similarities, particularly in with respect to possible “protective” factors, as well as important distinctions. Further study will be needed to determine if other cities, domestic or global, have comparable trends and hypotheses will need to be generated and tested to attempt to identify associations as more complete and reliable data become more available.

## Data Availability

All data used in this manuscript are from publicly available data sources.

